## Supplementary material for "Cumulative effects of particulate matter pollution and meteorological variables on the risk of influenza-like illness in Bialystok, Poland": Supplementary material (Tables S1,S2,S3).docx

Table S1. Reported influenza-like illness cases in Bialystok and Bialystok County divided by year and by age group.

| Age Year | 2013 | 2014 | 2015 | 2016 | 2017 | 2018 | 2019 | 2013-2019 |
| --- | --- | --- | --- | --- | --- | --- | --- | --- |
| 0-4 | 11,092 | 8,982 | 10,041 | 14,794 | 16,092 | 10,606 | 10,396 | 82,003 |
| 5-14 | 10,574 | 7,869 | 9,084 | 14,064 | 14,350 | 10,038 | 7,267 | 73,246 |
| 15-64 | 25,332 | 16,376 | 18,966 | 27,674 | 31,900 | 22,752 | 16,559 | 159,559 |
| 65+ | 4,716 | 2,563 | 3,224 | 4,975 | 6,226 | 5,711 | 3,764 | 31,179 |
| Total | 51,714 | 35,790 | 41,315 | 61,507 | 68,568 | 49,107 | 37,986 | 345,987 |

Table S2. Annual and 24-hour mean concentrations of particulate matter (µg/m^3^) in Bialystok.

| Year |  | Annual mean | SD | Min | P25 | Median | P75 | Max | n (%) > WHO daily  guidelines | n (%) missing |
| --- | --- | --- | --- | --- | --- | --- | --- | --- | --- | --- |
| 2013 | PM_2.5_ | 22.9 | 11.1 | 3.8 | 15.2 | 20.3 | 28.1 | 75.2 | 122 (35%) | 21 (6%) |
|  | PM_10_ | 23.2 | 13.7 | 3.9 | 13.9 | 20.3 | 28.8 | 96.5 | 13 (4%) | 0 |
| 2014 | PM_2.5_ | 24.9 | 11.7 | 5.9 | 16.0 | 22.9 | 32.8 | 69.7 | 151 (42%) | 4 (1%) |
|  | PM_10_ | 26.5 | 15.8 | 5.8 | 15.2 | 22.1 | 33.3 | 97.1 | 30 (8%) | 0 |
| 2015 | PM_2.5_ | 24.9 | 14.7 | 6.1 | 15.0 | 19.9 | 31.6 | 91.0 | 125 (36%) | 16 (5%) |
|  | PM_10_ | 25.3 | 18.1 | 4.7 | 13.1 | 19.6 | 30.4 | 115.4 | 40 (11%) | 0 |
| 2016 | PM_2.5_ | 19.8 | 9.1 | 3.3 | 12.9 | 18.1 | 25.2 | 65.2 | 77 (26%) | 65 (22%) |
|  | PM_10_ | 20.2 | 12.5 | 1.7 | 11.8 | 16.7 | 24.7 | 86.9 | 12 (3%) | 0 |
| 2017 | PM_2.5_ | 21.0 | 12.9 | 2.7 | 12.9 | 17.9 | 25.8 | 129.7 | 92 (27%) | 23 (7%) |
|  | PM_10_ | 20.2 | 14.4 | 2.7 | 11.5 | 17.0 | 24.2 | 139.1 | 12 (3%) | 0 |
| 2018 | PM_2.5_ | 24.0 | 13.3 | 2.4 | 14.2 | 21.4 | 29.9 | 88.4 | 132 (37%) | 9 (3%) |
|  | PM_10_ | 22.1 | 14.6 | 2.8 | 11.9 | 18.2 | 27.9 | 97.4 | 19 (5%) | 1 (<1%) |
| 2019 | PM_2.5_ | 19.0 | 9.3 | 2.9 | 12.8 | 17.6 | 22.3 | 64.4 | 68 (19%) | 3 (1%) |
|  | PM_10_ | 18.6 | 10.1 | 2.7 | 11.8 | 16.8 | 22.5 | 91.2 | 3 (1%) | 0 |

Table S3. Spearman’s correlation coefficients between the environmental factors and the incidence of influenza-like illness.

|  | Number of cases | Temperature (°C) | Relative humidity (%) | Absolute humidity (g/m^3^) | Wind speed (m/s) | Precipitation (mm) | Precipitation duration (hours) | Sunshine duration (hours) | PM_2.5_ (µg/m^3^) | PM_10_ (µg/m^3^) |
| --- | --- | --- | --- | --- | --- | --- | --- | --- | --- | --- |
| Number of cases | - | 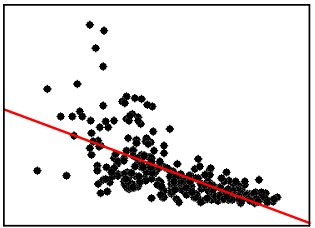 | 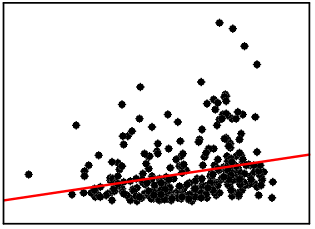 | 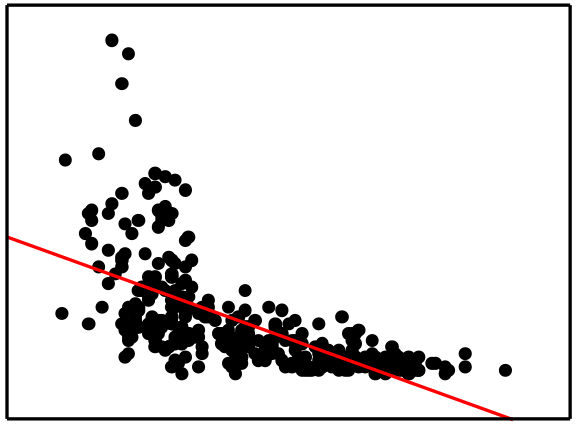 | 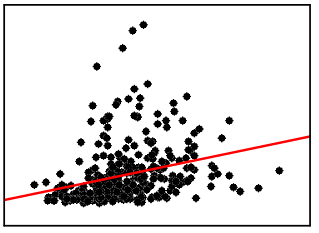 | 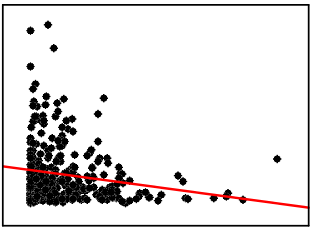 | 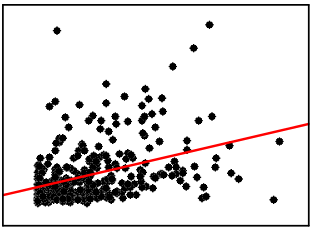 | 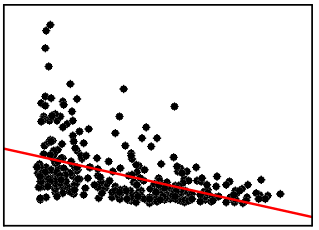 | 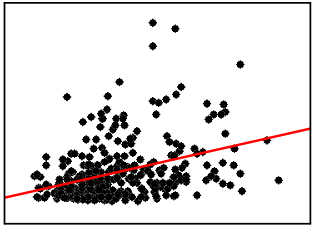 | 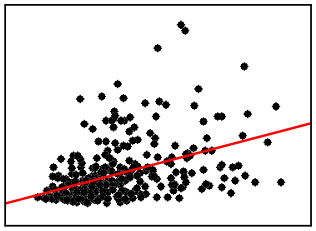 |
| Temperature (°C) | -0.78 *** | - | 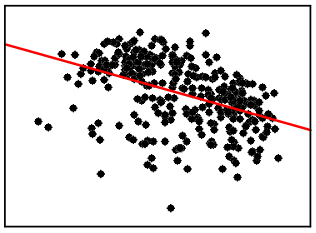 | 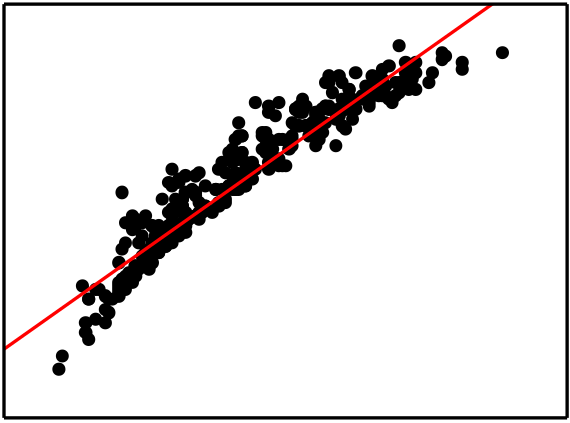 | 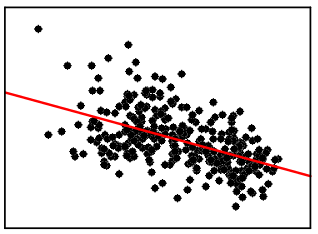 | 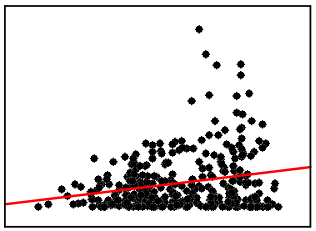 | 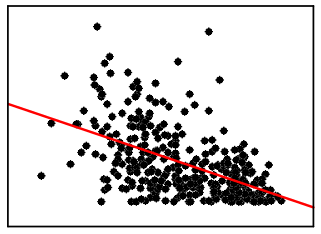 | 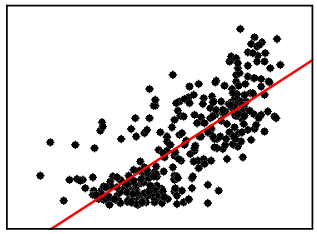 | 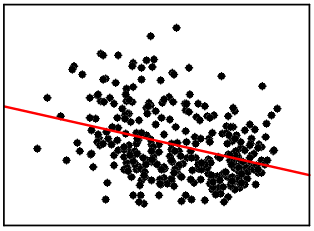 | 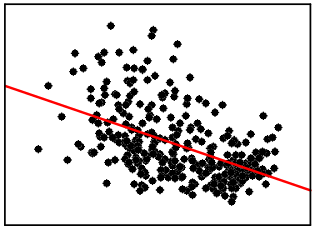 |
| Relative humidity (%) | 0.33 *** | -0.5 *** | - | 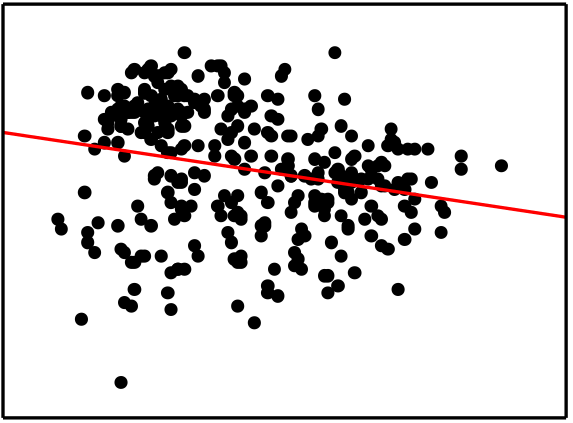 | 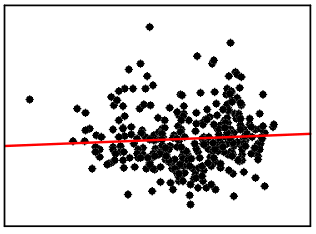 | 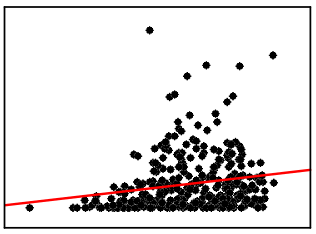 | 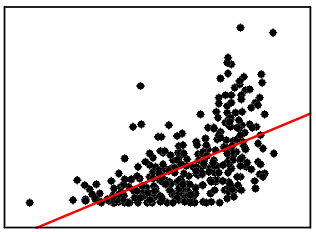 | 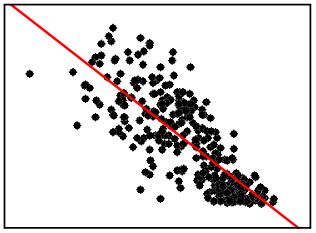 | 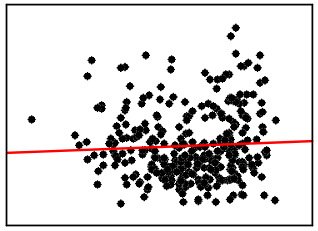 | 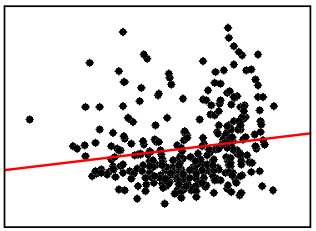 |
| Absolute humidity (g/m^3^) | -0,77 | 0,97 | -0,29 | - | 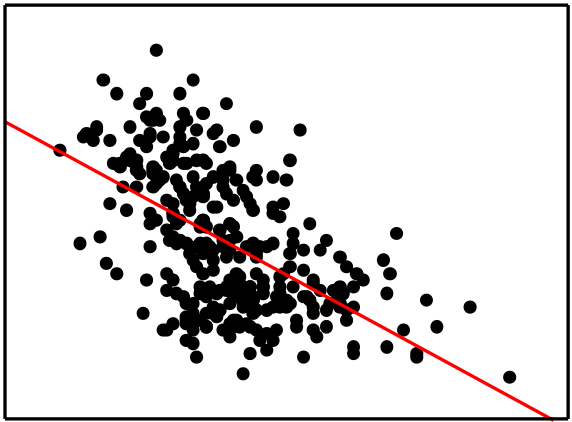 | 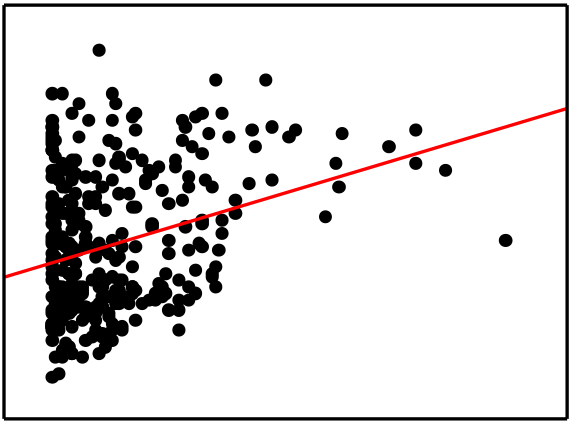 | 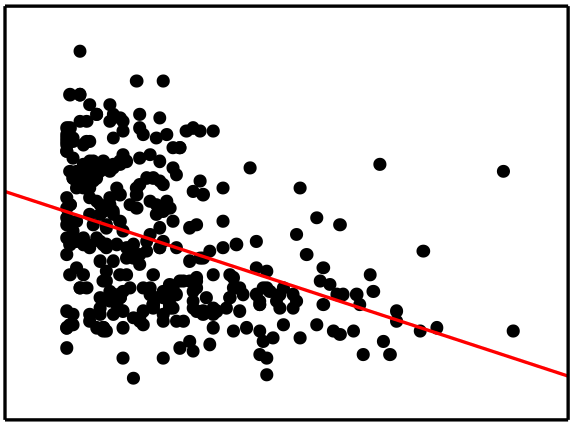 | 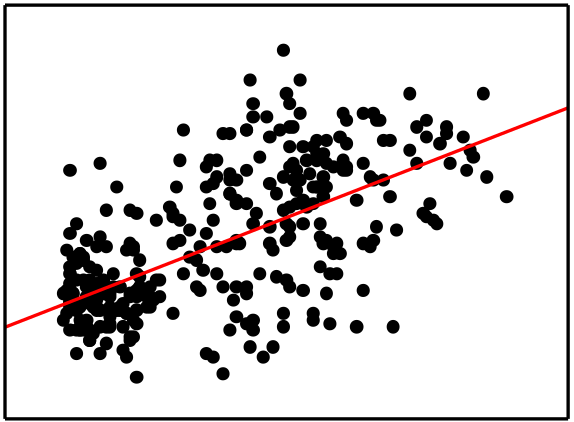 | 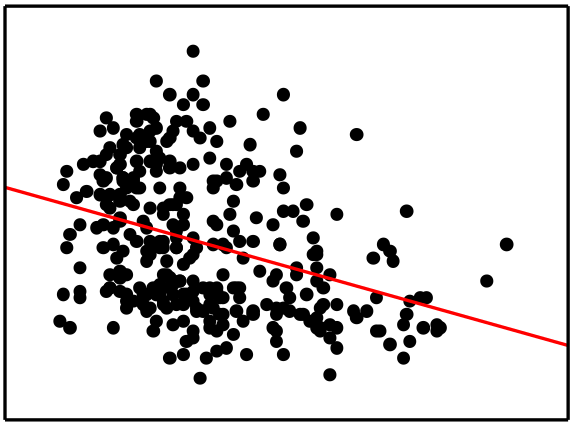 | 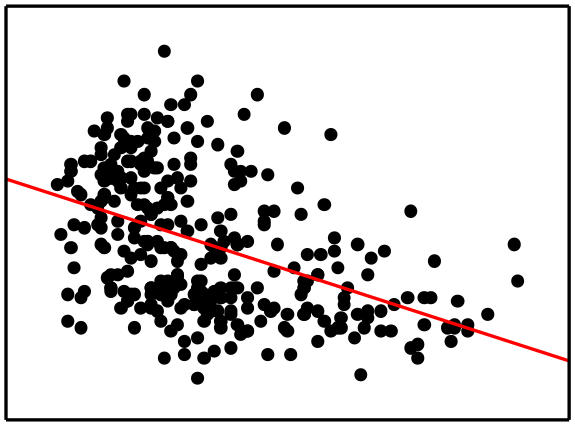 |
| Wind speed (m/s) | 0.43 *** | -0.52 *** | 0.13 * | -0,54 *** | - | 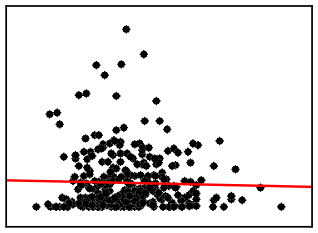 | 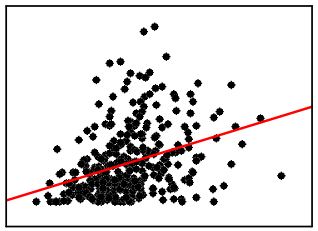 | 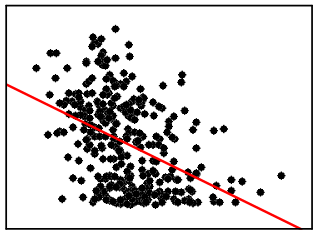 | 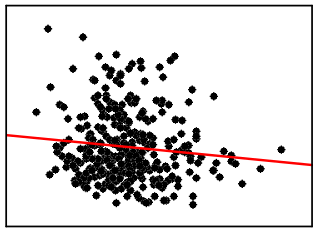 | 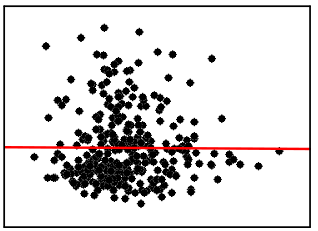 |
| Precipitation (mm) | -0.15 ** | 0.15 ** | 0.23 *** | 0,25 *** | *NS* | - | 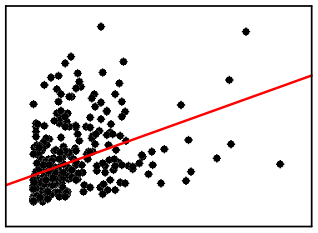 | 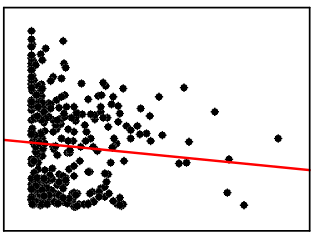 | 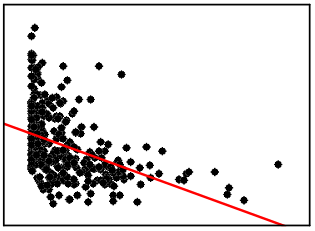 | 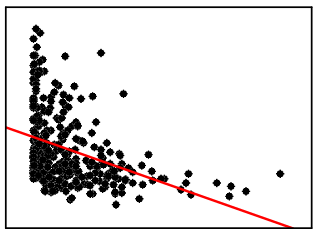 |
| Precipitation duration (hours) | 0.42 *** | -0.55 *** | 0.61 *** | -0,43 *** | 0.39 *** | 0.58 *** | - | 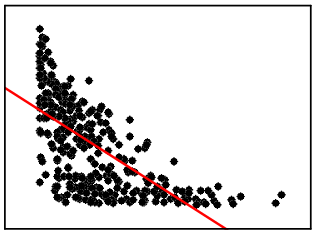 | 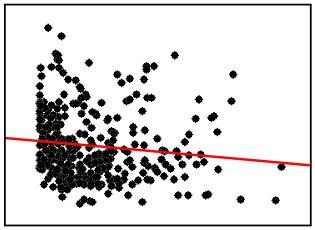 | 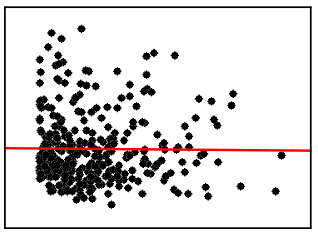 |
| Sunshine duration (hours) | -0.57 *** | 0.75 *** | -0.82 *** | 0,62 *** | -0.43 *** | -0.17 ** | -0.73 *** | - | 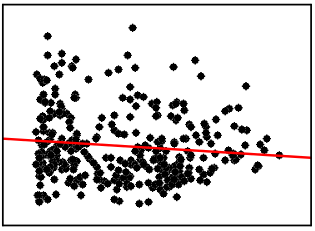 | 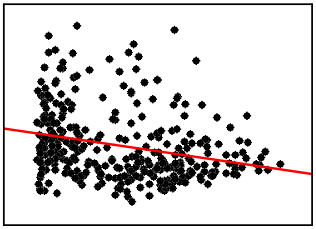 |
| PM_2.5_ (µg/m^3^) | 0.4 *** | -0.34 *** | *NS* | -0,37 *** | *NS* | -0.51 *** | -0.2 *** | *NS* | - | 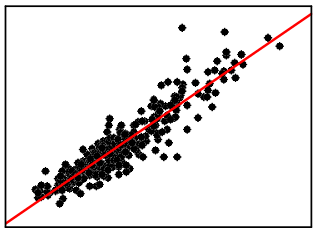 |
| PM_10_ (µg/m^3^) | 0.51 *** | -0.54 *** | 0.22 *** | -0,55 *** | *NS* | -0.46 *** | *NS* | -0.29 *** | 0.92 *** | - |

* indicates p < 0.05; ** indicates p < 0.01; *** indicates p < 0.001;
