## Supplementary figures and images for "Cumulative effects of particulate matter pollution and meteorological variables on the risk of influenza-like illness in Bialystok, Poland"

### Figure S1.pdf

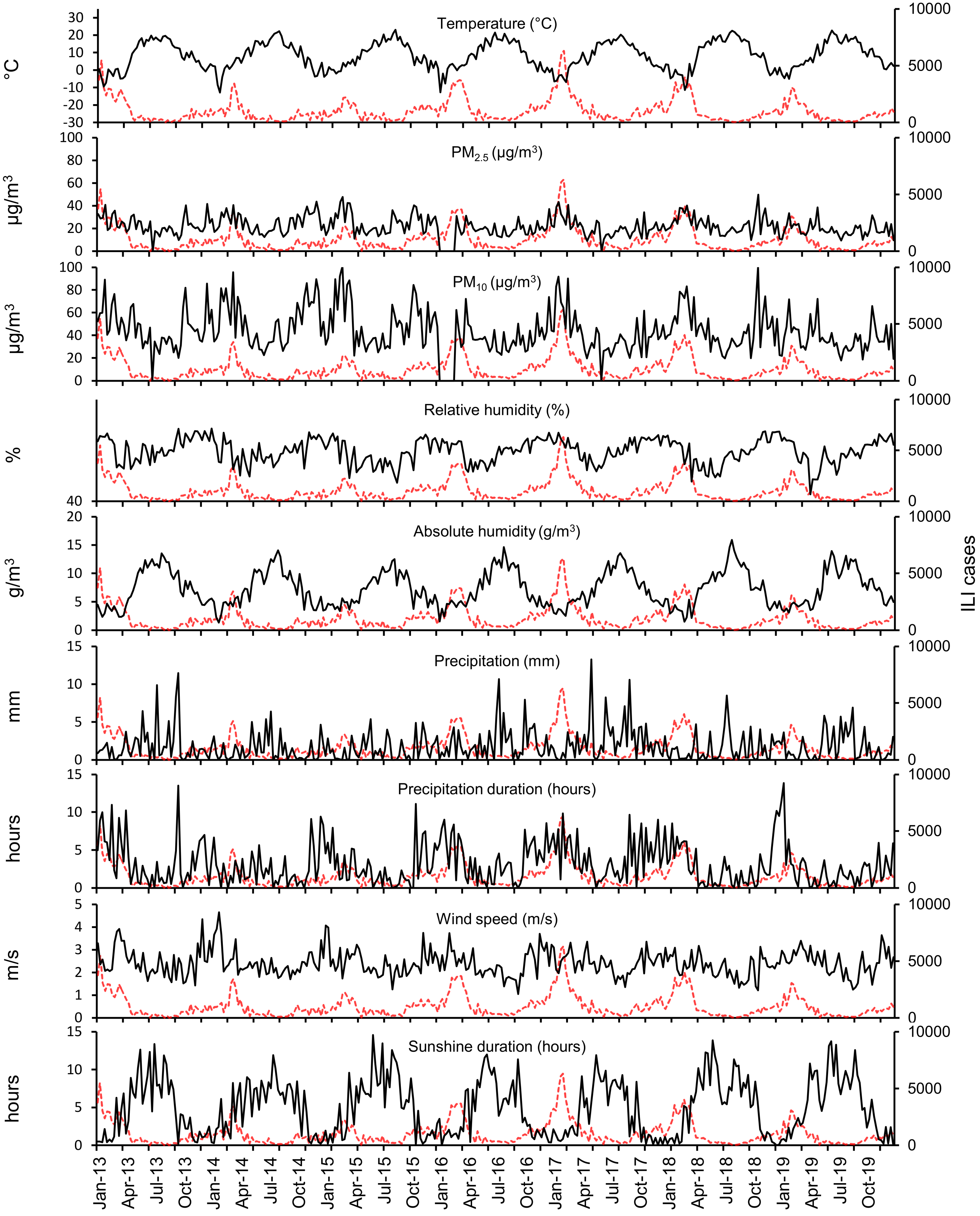

### Figure S2.tif

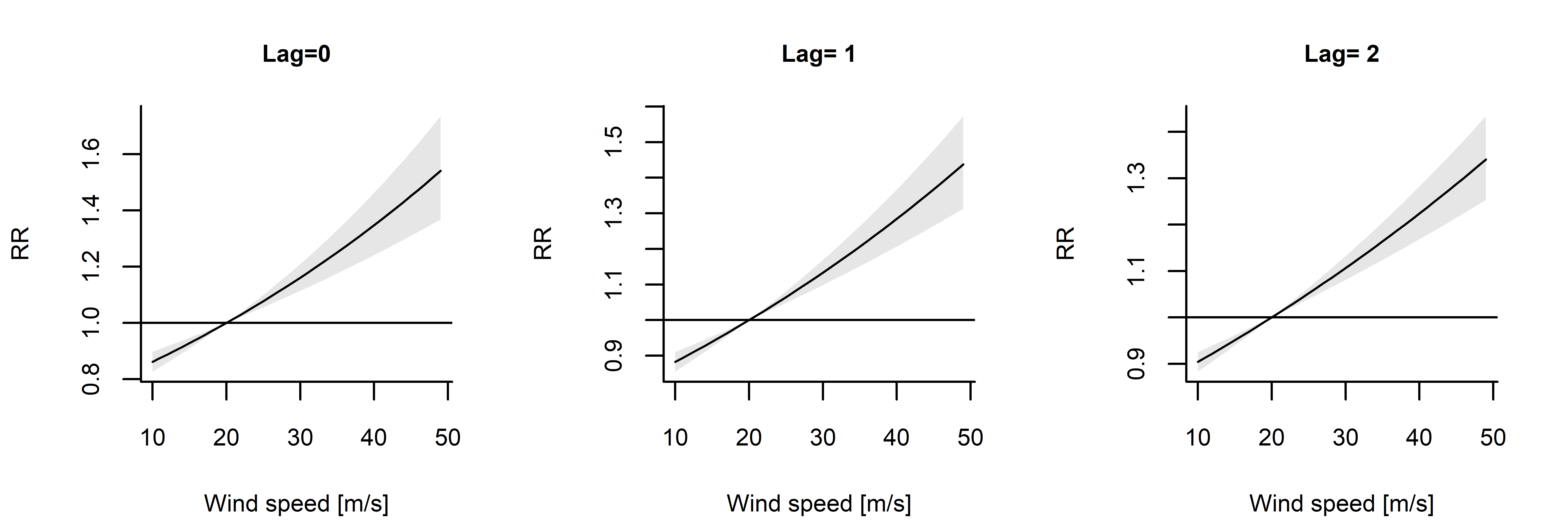
